# Systemic inflammation and endothelial dysfunction influence the risk and severity of hemorrhagic transformation after endovascular treatment for acute ischemic stroke

**DOI:** 10.64898/2026.05.08.26352571

**Authors:** Kattalin Alvarez Agoues, Patricia de la Riva, Jon Rodríguez-Antigüedad, Virginia Gomez, Gorka Arenaza, Ana Gorostidi, Noemí Díez, Ana de Arce, Maite Martínez Zabaleta, Eñaut Garmendia, Alex luttich, Maier Anabitarte, Jose Angel Larrea, Alberto Bergareche, Adolfo López de Munain, Juan Marta-Enguita

**Affiliations:** Neurology Department. Donostia University Hospital. San Sebastian. Spain; Ictus. Biogipuzkoa Health Research Institute. San Sebastian. Spain; Faculty of Health Sciences. Deusto University. San Sebastian. Spain; eurosciences Department. University of the Basque Country. San Sebastian. Spain; RICORS-ictus. National Institute of Health Carlos III. Madrid. Spain; Neurology Department. Hospital de la Santa Creu i Sant Pau. Barcelona. Spain; Radiology Department. Donostia University Hospital. San Sebastian. Spain; Genomics Platform. Biogipuzkoa Research Institute. San Sebastian. Spain; Neurodegenerative diseases. Biogipuzkoa Research Institute. San Sebastian. Spain; CIBERNED (CIBER, Institute Carlos III), Madrid. Spain; Valdecilla Health Research Institute. IDIVAL. Santander. Spain

**Keywords:** Acute ischemic stroke (AIS), Hemorrhagic transformation (HT), endovascular treatment (EVT), Neutrophil-to-lymphocyte ratio (NLR), Platelet-to-lymphocyte ratio (PLR), Systemic immune-inflammation index (SII), Endothelial dysfunction, Homoarginine (Harg), Flow-mediated dilation (FMD), Biomarkers

## Abstract

**Background:** Hemorrhagic transformation (HT) is a frequent and serious complication, occurring in up to 40% of cases after endovascular treatment (EVT) for acute ischemic stroke (AIS). Inflammation has been increasingly recognized as a key factor influencing both stroke pathophysiology and post-treatment complications (such as HT) interacting with endothelial dysfunction to exacerbate vascular injury after EVT. The objective of this study is to evaluate whether systemic inflammatory status predicts HT in AIS patients, and its relationship with endothelial biomarkers in the setting of this complication.

**Methods:** We retrospectively reviewed a prospective cohort of 229 AIS patients treated with EVT. Demographic, clinical, imaging, and laboratory data were collected. Inflammatory markers included white blood cell subsets and indices such as neutrophil-to-lymphocyte ratio (NLR), platelet-to-neutrophil ratio (PNR), systemic immune-inflammation index (SII), and systemic inflammation response index (SIRI). Endothelial function was assessed by flow-mediated dilation (FMD) and circulating homoarginine (HArg), asymmetric dimethylarginine (ADMA), and symmetric dimethylarginine (SDMA). The main outcome was radiological or symptomatic HT, classified according to ECASS criteria.

**Results:** HT was observed in 92 patients (40.2%), of whom 35 (36.1% of HT and 15.3% of the total) were symptomatic. In multivariate analysis, independent predictors of HT included higher NIHSS at admission, higher plasma glucose at admission, the use of non-aspiration devices, lower pre-recanalization lymphocyte count, higher pre-recanalization SII and higher NLR levels. Among endothelial function markers, HArg correlated with inflammatory markers, ANC (r = −0.2) and WBC (r = −0.19), and was associated to PH and symptomatic HT, but not with any radiologic HT after AIS.

**Conclusions:** An altered inflammatory status prior to EVT in AIS patients is associated with an increased risk of developing HT after EVT. Additionally, endothelial dysfunction could participate in the more aggressive forms of this complication.

## INTRODUCTION

Acute ischemic stroke (AIS) remains one of the most significant non-communicable diseases worldwide, standing as the second leading cause of mortality (approximately 7 million deaths) according to the Global Burden of Disease (GBD) 2021 report. [1] Currently, reperfusion therapy is the most effective treatment for AIS, which consists of intravenous thrombolysis (IVT) with tissue plasminogen activator (such as alteplase or tenecteplase) and endovascular therapy (EVT) in eligible patients. [2]

Hemorrhagic transformation (HT) is a major complication in the progression of AIS and a frequent adverse event following reperfusion therapies. Its incidence in follow-up neuroimaging is as high as 40% [3]. When HT leads to neurological deterioration, it is referred to as symptomatic intracranial hemorrhage (sICH), which is associated with high mortality and worse outcomes. [4]

Multiple factors have been associated with risk of HT following EVT, including, *inter alia*: epidemiological factors (age, pre-stroke treatment, previous diseases, etc.); characteristics of the infarct (size of ischemic core, severity, etc.); EVT treatment times; and poor collateral status[5]. However, the precise mechanisms underlying cerebral bleeding in individual patients are still poorly understood.

Identifying risk factors for HT is crucial to optimizing treatment strategies and minimizing adverse events. Among these potential determinants, inflammation has gained attention as a potential contributor to both ischemic injury and post-stroke complications. [6,7]. Inflammatory markers (such as C-reactive protein, calprotectin and interleukin-6) in peripheral blood have been investigated as potential indicators of stroke severity and prognosis [8] [9]. Recent studies have also explored the prognostic relevance of inflammatory indices, such as the neutrophil-to-lymphocyte ratio (NLR), the Systemic Inflammatory Index (SII), and the Systemic Inflammatory Response Index (SIRI). [6]

HT has also been linked to reperfusion injury in the setting of blood–brain barrier (BBB) disruption and impaired of cerebral blood flow autoregulation, processes in which the cerebrovascular endothelium plays a critical role through the synthesis of nitric oxide (NO), a powerful vasodilator. Genetic variations affecting the NO pathway has been associated with interindividual susceptibility to HT [10]. This endothelium-NO pathway is also known to interplay with inflammatory mediators from the earliest stages of the ischemic cascade in AIS and the roles of both pathophysiological pathways could be mutually enhanced to facilitate ischemia-reperfusion injury. [10,11]

Endothelial function can be evaluated through multiple approaches. One common approach is the measurement of flow-mediated dilation (FMD) via brachial ultrasound, which provides an assessment of systemic endothelial function[12]. Additionally, several NO-related metabolites, including arginine derivatives such as homoarginine (HArg), symmetric dimethylarginine (SDMA), and asymmetric dimethylarginine (ADMA), have been identified as meaningful biomarkers [13].

The aim of this study is to analyze whether patient’s inflammatory status before and after EVT is associated with the development of HT in AIS patients and to explore the interplay between inflammatory and endothelium biomarkers in this context.

## MATERIALS AND METHODS

The data supporting the study’s findings are available from the corresponding author upon request.

### Study Design

This is a retrospective review of a prospective cohort of patients treated with EVT and admitted to the Stroke Unit at Donostia University Hospital (DUH) between March 2018 and August 2020.

### Study Population

The study included patients diagnosed with AIS due to large vessel occlusion, as confirmed by emergency CT angiography (CTA), who underwent EVT at DUH during the period from March 2018 to August 2020.

The inclusion criteria for this study were as follows:

1. Patients presenting with a disabling focal neurological deficit and AIS with confirmed acute vessel occlusion on CTA, involving the proximal middle cerebral artery (MCA), the terminal internal carotid artery (TICA), or other large vessels including the anterior cerebral artery (ACA), posterior cerebral artery (PCA), basilar artery, vertebral artery, or extracranial carotid artery.
2. Availability of basal neuroimaging (CT and CTA) and a 24-hour follow-up CT scan to assess for HT after EVT.
3. Treatment with EVT, with or without prior intravenous thrombolysis (IVT), based on standard clinical criteria.

Between March 2018 and March 2020, patients were included consecutively in the study; however, from March 2020 to June 2020, due to the COVID19 outbreak, recruitment continued, but was non-consecutive.

### Descriptive Variables

The patients’ data were collected through medical record review and anonymized in a dedicated database.

The study included the following demographic variables: age, sex, and cardiovascular risk factors (previously known or diagnosed during admission), such as hypertension, diabetes mellitus, dyslipidemia, atrial fibrillation, prior stroke or transient ischemic attack, pre-stroke modified Rankin Scale (mRS), and prior use of antiplatelet and/or anticoagulant drugs.

Clinical variables included stroke severity by NIHSS (National Institutes of Health Stroke Scale) at admission, site of occlusion (e.g., internal carotid artery, segments of the MCA, ACA, PCA, basilar artery, vertebral artery, and extracranial carotid artery), and stroke etiology classified as atherothrombotic, cardioembolic, lacunar, or undetermined.

Variables related to reperfusion treatment included IVT, EVT device (aspiration, stent retriever, or combined technique), and time intervals such as onset-to-groin puncture, groin to reperfusion, door-to-reperfusion, and onset to reperfusion times. Recanalization success was assessed using the TICI (Thrombolysis in Cerebral Infarction) scale with grades ranging from 0 to 3.[14]Successful recanalization was defined as a TICI score of 2b, 2c, or 3, indicating reperfusion of more than 50% of the affected vascular territory. [15]

### Inflammatory variables

Inflammatory status assessment included pre- and post-recanalization measures of white blood cell count (WBC), absolute lymphocyte count (ALC), absolute neutrophil count (ANC), absolute monocyte count (AMC), and absolute platelet count (APC). Additionally, systemic inflammation markers such as C-reactive protein (CRP), the Systemic Immune-Inflammation Index (SII) and the Systemic Inflammation Response Index (SIRI) were defined as follows: SIRI = neutrophil count x monocyte count / lymphocyte count, SII = platelet count x neutrophil count/lymphocyte count.[6,16] Other indices, including the neutrophil-to-lymphocyte ratio (NLR) and neutrophil-to-platelet ratio (NPR), were also analyzed. NLR was calculated by dividing the neutrophil count by the lymphocyte count. NPR was calculated by dividing the neutrophil count by the platelet count. [17] [18]

For pre-EVT measures, venous blood samples were drawn from all patients in the emergency department before the initiation of any treatment as standard-of-care.

Biochemical parameters, including glucose levels, were analyzed. In addition, a complete blood count was performed to determine neutrophil, lymphocyte, monocyte, and platelet counts using standard laboratory techniques.

After receiving treatment, new blood samples were taken within the following 24 hours. Among other parameters, the following were measured: glucose, total cholesterol, low-density lipoprotein (LDL), high-density lipoprotein (HDL), triglycerides, and CRP. CRP was quantified using a particle-enhanced immunoturbidimetric assay with TRIS buffer and bovine serum albumin as reagent, performed on a Cobas c 701/702 analyzer (Roche Diagnostics, Germany). The remaining biochemical parameters were determined with specific enzymatic/colorimetric kits on the same analyzer. A repeat complete blood count was performed to determine neutrophil, lymphocyte, monocyte, and platelet counts, and coagulation parameters were also reassessed.

In addition, serum samples were obtained from arterial blood extracted immediately following femoral artery puncture during EVT. ADMA, SDMA and HArg, were quantified in these samples and in conditioned cell culture media using an ultra-performance liquid chromatography–tandem mass spectrometry method as described in detail in **Supplementary Methods**. The laboratory performing and analyzing the mass spectrometric measurements was blinded to the clinical status of the patients.

### Outcome variables

The main outcome was HT, defined according to the European Cooperative Acute Stroke Study (ECASS) radiological classification: hemorrhagic infarction (HI) type 1, isolated petechiae without mass effect; HI type 2, confluent petechiae without mass effect; parenchymal hemorrhage (PH) type 1, homogeneous hyperdensity with minimal mass effect involving <30% of the infarcted area; PH type 2, hyperdensity involving >30% of the infarcted area with definite mass effect, possible intraventricular extension, or hemorrhage outside the infarcted area[20]. Secondary outcomes were: Modified Rankin Scale (mRS) at discharge and at 90 days post-stroke, good clinical outcome (GCO) (defined as a mRS equal or less than 2) at discharge and at 90 days, futile recanalization (defined as poor 90 days functional status (mRs ≥3) despite complete or near-complete recanalization, TICI ≥2b)[6].

Symptomatic HT (sHT) was considered as any HT that worsened by ≥4 points on the NIHSS according to the ECASS-II definition. [21]

### Statistical Analysis

Categorical variables are presented as the number and percentage (%) of cases, while continuous quantitative variables are expressed using their mean and standard deviation (SD) or median and interquartile range (IQR), as appropriate. Normality distribution of the data was assessed using the Kolmogorov-Smirnov test. Variables that did not follow a normal distribution were log-transformed and their distribution was reevaluated before being included in adjusted models. First, the study population was divided into two groups: “HT” (the group with any type of radiological HT) and “no HT”. Within the HT group, a distinction was also made regarding whether the HT was symptomatic or not, and each HT was further classified into ECASS radiological subgroups. Baseline characteristics of the study groups were compared using the Chi-square test for categorical variables, the Student’s T-test for quantitative variables with a normal distribution, and the Mann-Whitney U test for quantitative variables without a normal distribution, ordinal variables or when the assumption of homogeneity of variance between the groups is violated. The relationships between endothelial function and inflammatory markers were assessed using Pearson correlation coefficients.

Independent predictors of HT were identified through binary and multivariate logistic regression analyses, including variables with p < 0.05 in the univariate analysis. In a second model, sex and age were forced in addition to significant univariate predictors. To minimize collinearity, one variable was excluded when two were highly correlated. Given the expected intercorrelations among inflammatory and endothelial markers, these variables were entered separately into the multivariate model if appropriate. Arginine is listed as a component of the alteplase commercial product, the compound used for IVT in this study, and thus, patients receiving IVT are expected to have higher levels of HArg. To control for this possible confounding factor, we decided to include IVT in the multivariate regression analysis if HArg was included.

Statistical analysis was conducted using IBM SPSS Statistics, version 26 (Chicago, Illinois, USA). Statistical significance was set at p < 0.05

### Ethical Considerations

The study was approved by the Ethics Committee of Gipuzkoa on February 20, 2018 under protocol code PRI-CII-2018-01. Written informed consent was obtained from all patients in the study or their relatives, who gave permission to include their data in the study database in accordance with the Spanish Personal Data Protection law.

## RESULTS

During the study period, 233 patients with ischemic stroke underwent EVT. Two were excluded due to missing laboratory data, one due to abnormal results related to myelodysplastic syndrome, and one due to failure of recanalization treatment caused by vascular access difficulties. Ultimately, 229 patients were included. Baseline characteristics are presented in **Table 1**.

**Table 1.**
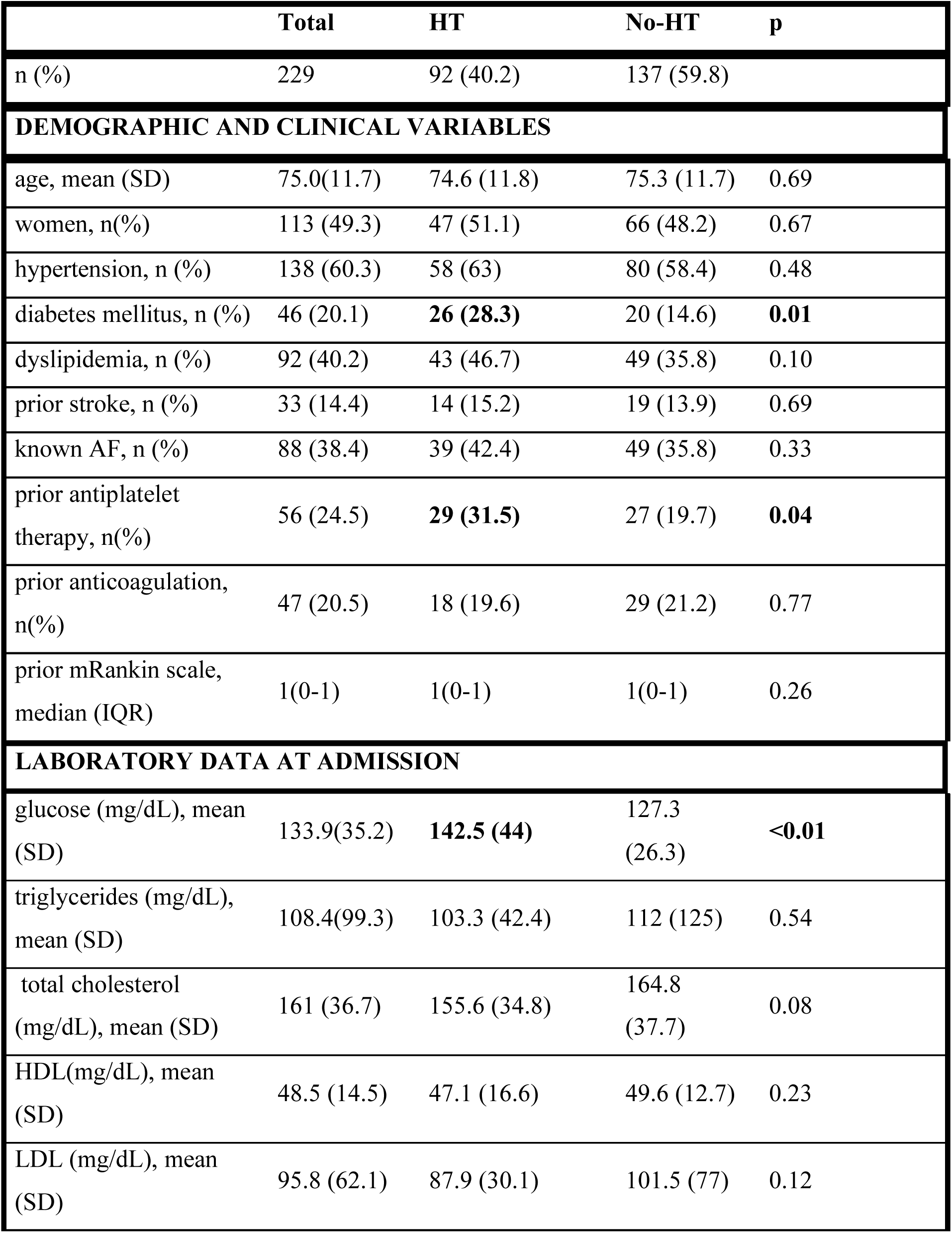
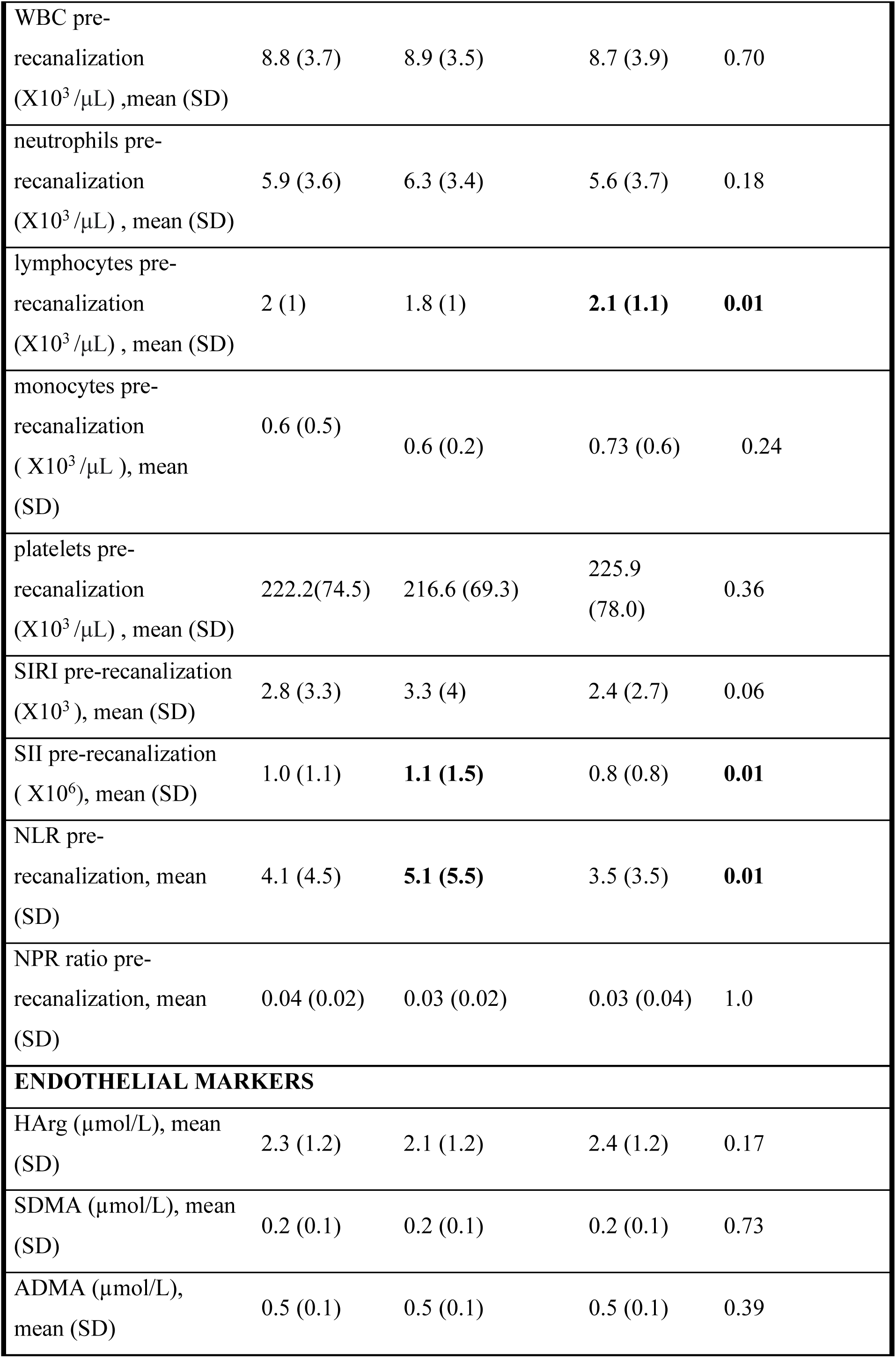
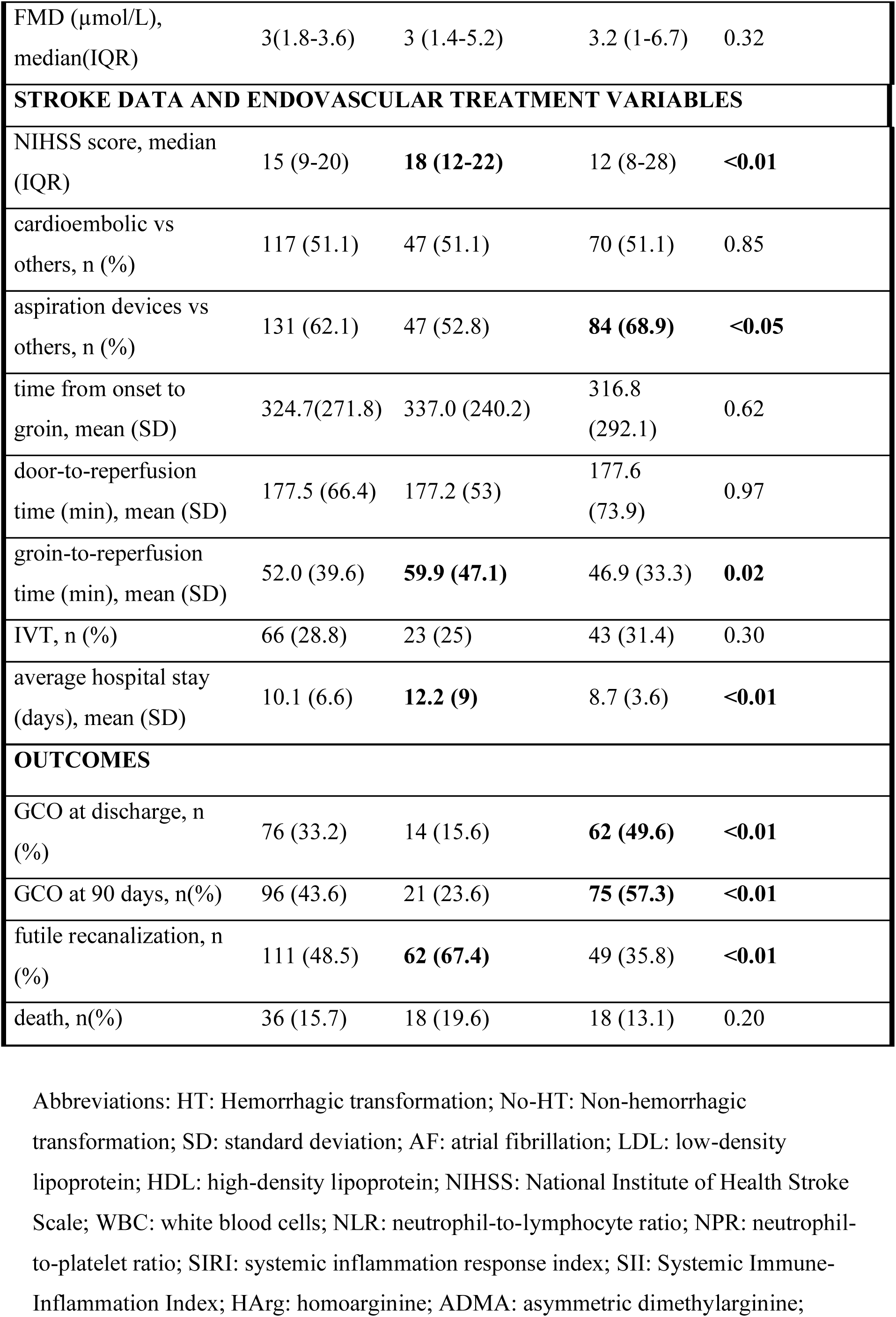
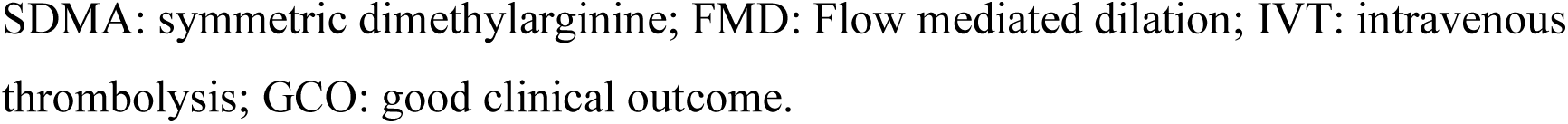
Univariate analysis and comparison of demographic, clinical, laboratory, and endovascular treatment variables between groups.

|  | Total | HT | No-HT | p |
| --- | --- | --- | --- | --- |
| n (%) | 229 | 92 (40.2) | 137 (59.8) |  |
| **DEMOGRAPHIC AND CLINICAL VARIABLES** |  |  |  |  |
| age, mean (SD) | 75.0(11.7) | 74.6 (11.8) | 75.3 (11.7) | 0.69 |
| women, n(%) | 113 (49.3) | 47 (51.1) | 66 (48.2) | 0.67 |
| hypertension, n (%) | 138 (60.3) | 58 (63) | 80 (58.4) | 0.48 |
| diabetes mellitus, n (%) | 46 (20.1) | **26 (28.3)** | 20 (14.6) | **0.01** |
| dyslipidemia, n (%) | 92 (40.2) | 43 (46.7) | 49 (35.8) | 0.10 |
| prior stroke, n (%) | 33 (14.4) | 14 (15.2) | 19 (13.9) | 0.69 |
| known AF, n (%) | 88 (38.4) | 39 (42.4) | 49 (35.8) | 0.33 |
| prior antiplatelet therapy, n(%) | 56 (24.5) | **29 (31.5)** | 27 (19.7) | **0.04** |
| prior anticoagulation, n(%) | 47 (20.5) | 18 (19.6) | 29 (21.2) | 0.77 |
| prior mRankin scale, median (IQR) | 1(0-1) | 1(0-1) | 1(0-1) | 0.26 |
| **LABORATORY DATA AT ADMISSION** |  |  |  |  |
| glucose (mg/dL), mean (SD) | 133.9(35.2) | **142.5 (44)** | 127.3 (26.3) | **<0.01** |
| triglycerides (mg/dL), mean (SD) | 108.4(99.3) | 103.3 (42.4) | 112 (125) | 0.54 |
| total cholesterol (mg/dL), mean (SD) | 161 (36.7) | 155.6 (34.8) | 164.8 (37.7) | 0.08 |
| HDL(mg/dL), mean (SD) | 48.5 (14.5) | 47.1 (16.6) | 49.6 (12.7) | 0.23 |
| LDL (mg/dL), mean (SD) | 95.8 (62.1) | 87.9 (30.1) | 101.5 (77) | 0.12 |
| WBC pre-recanalization<br>(X10<sup>3</sup> /μL) ,mean (SD) | 8.8 (3.7) | 8.9 (3.5) | 8.7 (3.9) | 0.70 |
| neutrophils pre-recanalization<br>(X10<sup>3</sup> /μL) , mean (SD) | 5.9 (3.6) | 6.3 (3.4) | 5.6 (3.7) | 0.18 |
| lymphocytes pre-recanalization<br>(X10<sup>3</sup> /μL) , mean (SD) | 2 (1) | 1.8 (1) | **2.1 (1.1)** | **0.01** |
| monocytes pre-recanalization<br>( X10<sup>3</sup> /μL ), mean (SD) | 0.6 (0.5) | 0.6 (0.2) | 0.73 (0.6) | 0.24 |
| platelets pre-recanalization<br>(X10<sup>3</sup> /μL) , mean (SD) | 222.2(74.5) | 216.6 (69.3) | 225.9 (78.0) | 0.36 |
| SIRI pre-recanalization<br>(X10<sup>3</sup> ), mean (SD) | 2.8 (3.3) | 3.3 (4) | 2.4 (2.7) | 0.06 |
| SII pre-recanalization<br>( X10<sup>6</sup>), mean (SD) | 1.0 (1.1) | **1.1 (1.5)** | 0.8 (0.8) | **0.01** |
| NLR pre-recanalization, mean<br>(SD) | 4.1 (4.5) | **5.1 (5.5)** | 3.5 (3.5) | **0.01** |
| NPR ratio pre-recanalization, mean<br>(SD) | 0.04 (0.02) | 0.03 (0.02) | 0.03 (0.04) | 1.0 |
| **ENDOTHELIAL MARKERS** |  |  |  |  |
| HArg (μmol/L), mean<br>(SD) | 2.3 (1.2) | 2.1 (1.2) | 2.4 (1.2) | 0.17 |
| SDMA (μmol/L), mean<br>(SD) | 0.2 (0.1) | 0.2 (0.1) | 0.2 (0.1) | 0.73 |
| ADMA (μmol/L),<br>mean (SD) | 0.5 (0.1) | 0.5 (0.1) | 0.5 (0.1) | 0.39 |
| FMD (μmol/L),<br>median(IQR) | 3(1.8-3.6) | 3 (1.4-5.2) | 3.2 (1-6.7) | 0.32 |
| **STROKE DATA AND ENDOVASCULAR TREATMENT VARIABLES** |  |  |  |  |
| NIHSS score, median<br>(IQR) | 15 (9-20) | **18 (12-22)** | 12 (8-28) | **<0.01** |
| cardioembolic vs<br>others, n (%) | 117 (51.1) | 47 (51.1) | 70 (51.1) | 0.85 |
| aspiration devices vs<br>others, n (%) | 131 (62.1) | 47 (52.8) | **84 (68.9)** | **<0.05** |
| time from onset to<br>groin, mean (SD) | 324.7(271.8) | 337.0 (240.2) | 316.8<br>(292.1) | 0.62 |
| door-to-reperfusion<br>time (min), mean (SD) | 177.5 (66.4) | 177.2 (53) | 177.6<br>(73.9) | 0.97 |
| groin-to-reperfusion<br>time (min), mean (SD) | 52.0 (39.6) | **59.9 (47.1)** | 46.9 (33.3) | **0.02** |
| IVT, n (%) | 66 (28.8) | 23 (25) | 43 (31.4) | 0.30 |
| average hospital stay<br>(days), mean (SD) | 10.1 (6.6) | **12.2 (9)** | 8.7 (3.6) | **<0.01** |
| **OUTCOMES** |  |  |  |  |
| GCO at discharge, n<br>(%) | 76 (33.2) | 14 (15.6) | **62 (49.6)** | **<0.01** |
| GCO at 90 days, n(%) | 96 (43.6) | 21 (23.6) | **75 (57.3)** | **<0.01** |
| futile recanalization, n<br>(%) | 111 (48.5) | **62 (67.4)** | 49 (35.8) | **<0.01** |
| death, n(%) | 36 (15.7) | 18 (19.6) | 18 (13.1) | 0.20 |

Abbreviations: HT: Hemorrhagic transformation; No-HT: Non-hemorrhagic transformation; SD: standard deviation; AF: atrial fibrillation; LDL: low-density lipoprotein; HDL: high-density lipoprotein; NIHSS: National Institute of Health Stroke Scale; WBC: white blood cells; NLR: neutrophil-to-lymphocyte ratio; NPR: neutrophil-to-platelet ratio; SIRS: systemic inflammation response index; SII: Systemic Immune-Inflammation Index; HArg: homoarginine; ADMA: asymmetric dimethylarginine;
SDMA: symmetric dimethylarginine; FMD: Flow mediated dilation; IVT: intravenous thrombolysis; GCO: good clinical outcome.

The mean age of the participants was 75.03 years (SD = 11.7), with 116 male patients (50.7%). Upon admission, the NIHSS median score was 15 (IQR = 11). The occluded artery was distributed as follows: 33 TICA (14.4%), 112 MCA1 (48.9%), 54 MCA2 (23.6%), 5 isolated extracranial carotid artery (2.2%), 2 ACA (0.8%), 3 PCA (1.3%), 13 Basilar artery and (5.7%) and 7 vertebral arteries (3.1%). At discharge, 76 patients (33.2%) were independent in their activities of daily living. This percentage increased to 96 patients (43.6%) at 3 months post-discharge.

For the following analysis, patients were divided into two groups: 92 patients (40.2%) who suffered a radiological HT on follow-up neuroimaging were assigned to the “HT” group, and 137 patients (59.8%) were assigned to the “No HT” group. On the other hand, symptomatic HT was present in 35 patients (15.3%).

Regarding the specific radiologic HT subtypes across the entire study population, 27 patients (11.8%) presented with HI1, 24 patients (10.5%) with HI2, 16 patients (7.0%) with PH1, and 23 patients (10.0%) with PH2. Therefore, parenchymal hemorrhage was observed in 39 out of the 92 patients with HT (42.4%).

Patients in the HT group were more likely to have been treated with antiplatelet therapy (31.5% vs 19.7%, p=0.04), and presented with more severe strokes upon admission, with a median NIHSS score 6 points higher (p<0.01). Additionally, patients in the HT group had a higher mean plasma glucose level upon admission (142.5 vs. 127.3, p<0.01), with a greater proportion of diabetic patients (56.5% vs 43.5%, p=0.01).

Regarding prognostic variables, the average hospital stay was 3,5 days longer in the HT group (12.2 vs. 8.7, p<0.05), and there were significantly fewer patients with GCO at discharge (15.6% vs 49.6%, p<0.01) and at 90 days post-discharge (23.6% vs 57.3%, p<0.01) without significant differences in mortality. Furthermore, a higher proportion of patients in the HT group experienced futile recanalization (67.4% vs. 35.8%, p < 0.01) compared to the No-HT group.

### Inflammatory Markers

Mean time from onset to blood sample extraction in the emergency department (pre-treatment) was 204.5 minutes (SD=199.5).

Regarding inflammatory parameters (**Table 1**), some differences were observed between both groups at admission. Lymphopenia was observed more frequently in patients in the HT group (ALC: 1.8 vs. 2.2; p=0.01). Meanwhile, the SII and the mean NLR were higher in the HT group (SII: 1.2 vs. 0.8; p=0.03) (NLR: 5.2 vs. 3.5; p=0.02).

The univariate analysis did not reveal significant differences post-recanalization between the two groups for the analyzed inflammatory markers (**Supplementary table 1**)

The multivariate analysis (**Table 2**) identified several independent predictors of HT. Stroke severity, reflected by a higher NIHSS at admission [OR (95%CI): 1.11 (1.05-1.17); p<0.01], and higher plasma glucose levels at admission [per each 10mg increase (95%CI): 1.17 (1.05-1.32); p<0.05] showed a strong association with HT. On the other hand, the use of aspiration devices [OR (95%CI): 0.38 (0.18-0.77); p<0.01] was associated with a lower risk of HT. Among inflammatory parameters, lower pre-recanalization ALC [per each 10^3^ increases: OR (95%CI): 0.96 (0.93-0.99); p<0.05], higher pre-recanalization SII [per each 10^6^ increase: 1.35 (1.02-1.80); p<0.05] and higher NLR levels [OR (95%CI): 1.11 (1.02-1.22; p<0.05] were associated to the risk of HT (**Table 2**).

**Table 2.**
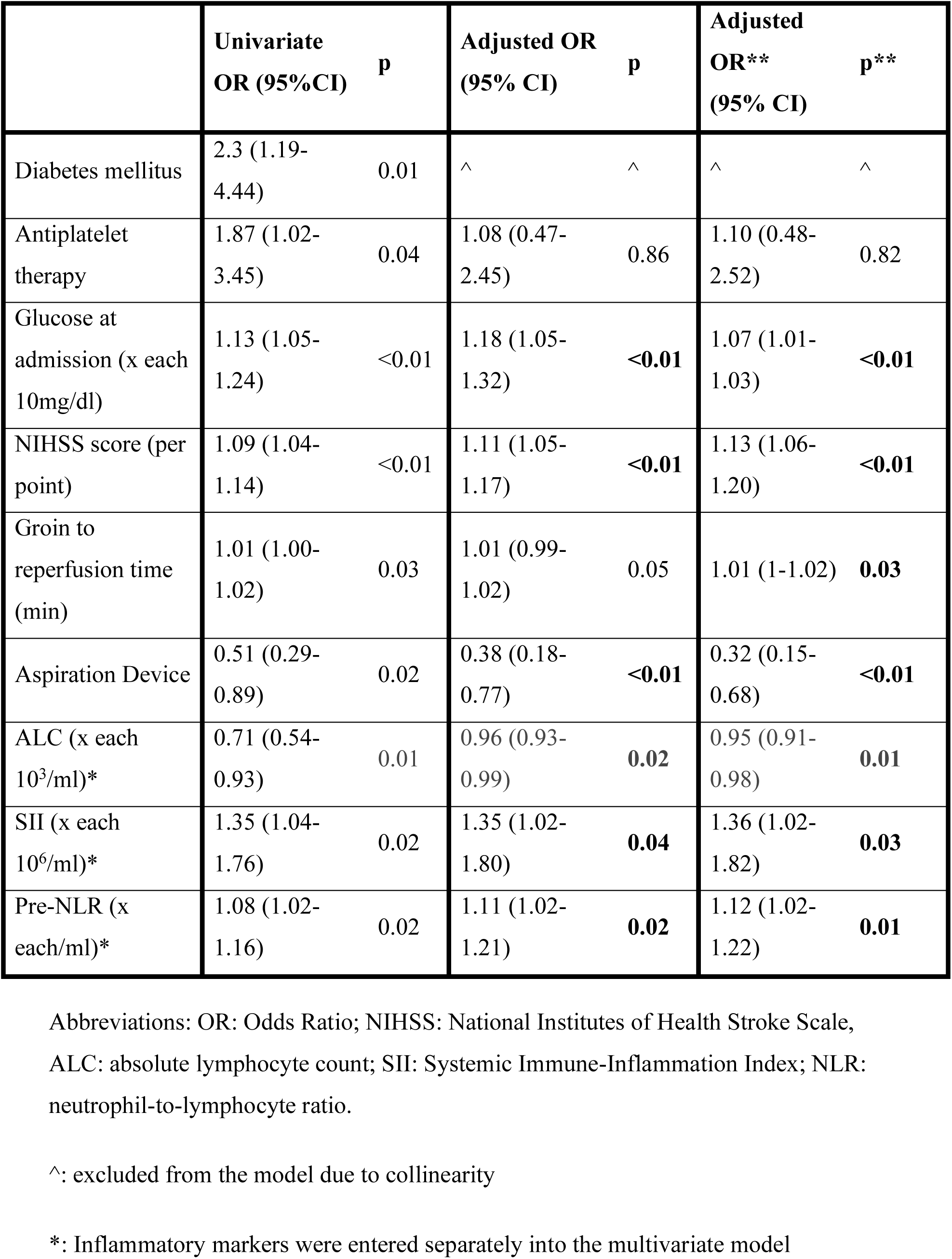
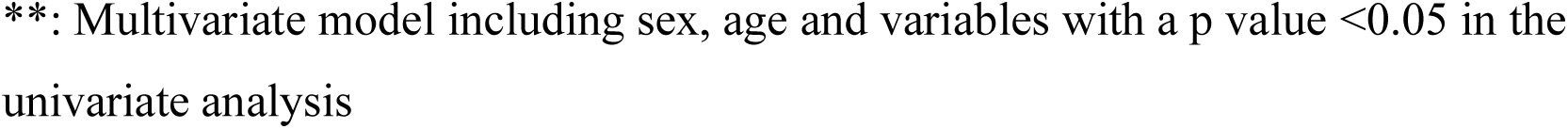
Univariate and multivariate logistic regression analysis of factors influencing HT after stroke treated with EVT.

|  | Univariate<br>OR (95%CI) | p | Adjusted OR<br>(95% CI) | p | Adjusted<br>OR**<br>(95% CI) | p** |
| --- | --- | --- | --- | --- | --- | --- |
| Diabetes mellitus | 2.3 (1.19-4.44) | 0.01 | ^ | ^ | ^ | ^ |
| Antiplatelet therapy | 1.87 (1.02-3.45) | 0.04 | 1.08 (0.47-2.45) | 0.86 | 1.10 (0.48-2.52) | 0.82 |
| Glucose at admission (x each 10mg/dl) | 1.13 (1.05-1.24) | <0.01 | 1.18 (1.05-1.32) | <0.01 | 1.07 (1.01-1.03) | <0.01 |
| NIHSS score (per point) | 1.09 (1.04-1.14) | <0.01 | 1.11 (1.05-1.17) | <0.01 | 1.13 (1.06-1.20) | <0.01 |
| Groin to reperfusion time (min) | 1.01 (1.00-1.02) | 0.03 | 1.01 (0.99-1.02) | 0.05 | 1.01 (1-1.02) | **0.03** |
| Aspiration Device | 0.51 (0.29-0.89) | 0.02 | 0.38 (0.18-0.77) | <0.01 | 0.32 (0.15-0.68) | <0.01 |
| ALC (x each 10<sup>3</sup>/ml)* | 0.71 (0.54-0.93) | 0.01 | 0.96 (0.93-0.99) | **0.02** | 0.95 (0.91-0.98) | **0.01** |
| SII (x each 10<sup>6</sup>/ml)* | 1.35 (1.04-1.76) | 0.02 | 1.35 (1.02-1.80) | **0.04** | 1.36 (1.02-1.82) | **0.03** |
| Pre-NLR (x each/ml)* | 1.08 (1.02-1.16) | 0.02 | 1.11 (1.02-1.21) | **0.02** | 1.12 (1.02-1.22) | **0.01** |

Abbreviations: OR: Odds Ratio; NIHSS: National Institutes of Health Stroke Scale, ALC: absolute lymphocyte count; SII: Systemic Immune-Inflammation Index; NLR: neutrophil-to-lymphocyte ratio.
^: excluded from the model due to collinearity
\*: Inflammatory markers were entered separately into the multivariate model
**\*\***: Multivariate model including sex, age and variables with a p value $<0.05$ in the univariate analysis

### Endothelial function and inflammatory markers

FMD was performed in 167 patients (72.9%) and the levels of plasma endothelial markers (HArg, SDMA and ADMA) were measured in 148 (64.6%) of the patients. Losses were due to technical issues and the COVID pandemia period where blood samples could not be processed. The time from stroke onset to arterial blood sample collection was 244.5 minutes (SD 195).

Arginine derivatives showed significant correlations with one another but not with FMD (**Supplementary table 2**). Correlation analysis between endothelial markers and inflammatory markers revealed pretreatment WBC and ANC correlated with HArg and SDMA correlated with preALC. No correlation was found between systemic endothelial function assessed by FMD and inflammatory markers (**Figure 1**).

Regarding HT, endothelial markers did not show significant association with radiological HT (**Table 1**) whereas HArg levels (**Figure 2**) were independently associated with the radiological PH subtype of HT in a univariate analysis (2.43 vs 1.79, p<0.01) and in a multivariate model including the variables associated with any HT (glucose at admission, NIHSS score, aspiration device) [aOR (95%CI): 0.53 (0.3-0.9), p=0.03]. In this context, when assessing symptomatic HT, we found that patients with sHT exhibited lower pre-recanalization HArg levels [1.8 vs 2.4, p<0.05] and independently predicted sHT (aOR(95%CI): 0.46 (0.22-0.95), p=0.04) in the multivariate analysis **(See main tables 3 and 4).**

**Table 3.**
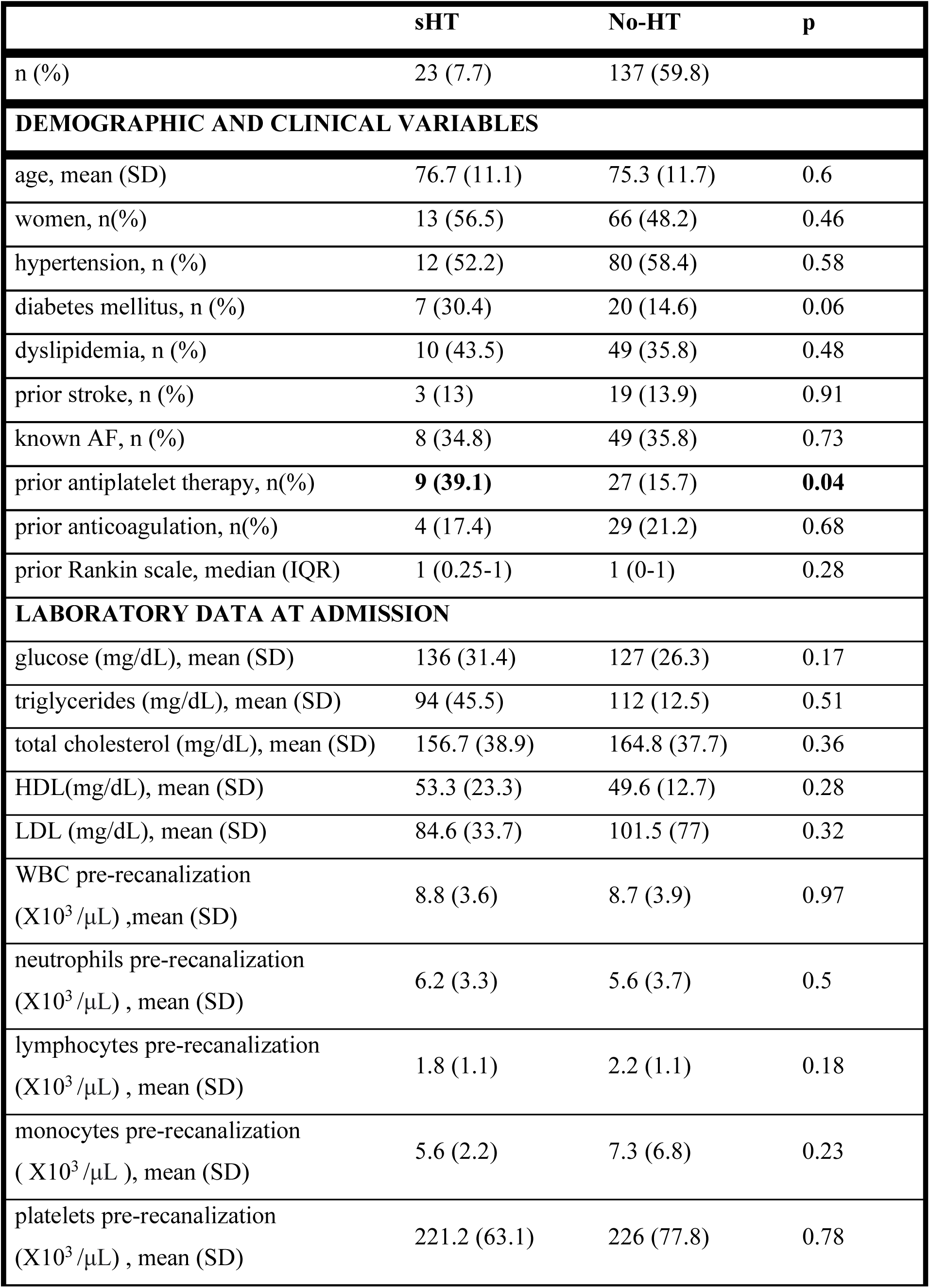
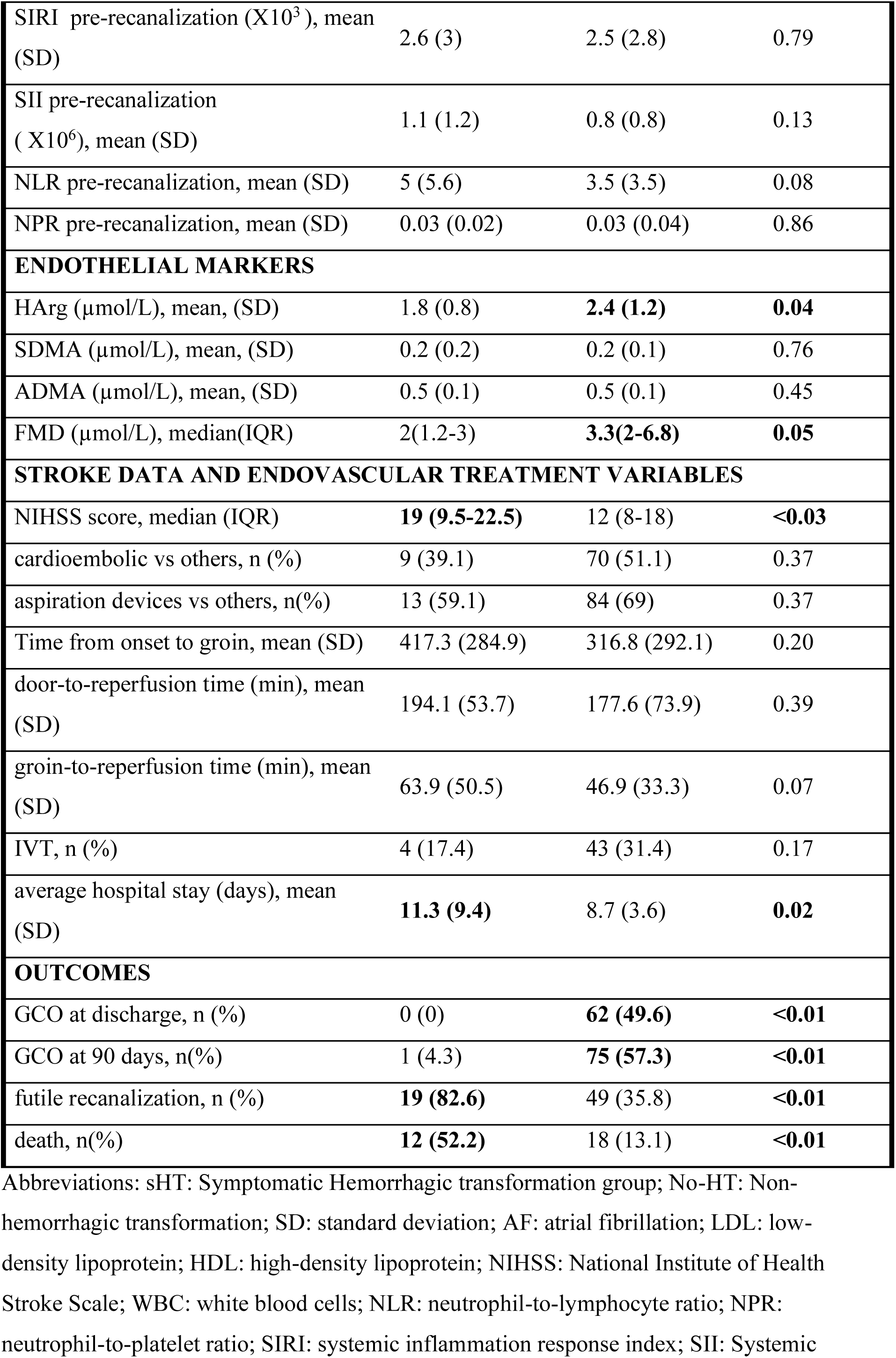
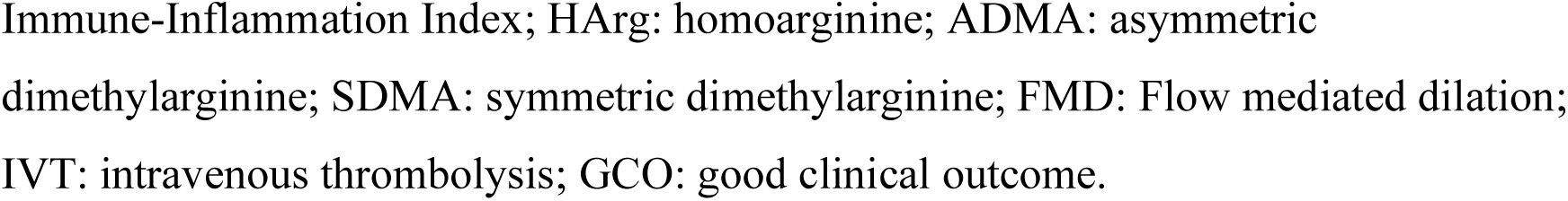
Comparison of demographic, clinical, laboratory, and endovascular treatment variables between symptomatic HT and No-HT patients.

|  | sHT | No-HT | p |
| --- | --- | --- | --- |
| n (%) | 23 (7.7) | 137 (59.8) |  |
| **DEMOGRAPHIC AND CLINICAL VARIABLES** |  |  |  |
| age, mean (SD) | 76.7 (11.1) | 75.3 (11.7) | 0.6 |
| women, n(%) | 13 (56.5) | 66 (48.2) | 0.46 |
| hypertension, n (%) | 12 (52.2) | 80 (58.4) | 0.58 |
| diabetes mellitus, n (%) | 7 (30.4) | 20 (14.6) | 0.06 |
| dyslipidemia, n (%) | 10 (43.5) | 49 (35.8) | 0.48 |
| prior stroke, n (%) | 3 (13) | 19 (13.9) | 0.91 |
| known AF, n (%) | 8 (34.8) | 49 (35.8) | 0.73 |
| prior antiplatelet therapy, n(%) | **9 (39.1)** | 27 (15.7) | **0.04** |
| prior anticoagulation, n(%) | 4 (17.4) | 29 (21.2) | 0.68 |
| prior Rankin scale, median (IQR) | 1 (0.25-1) | 1 (0-1) | 0.28 |
| **LABORATORY DATA AT ADMISSION** |  |  |  |
| glucose (mg/dL), mean (SD) | 136 (31.4) | 127 (26.3) | 0.17 |
| triglycerides (mg/dL), mean (SD) | 94 (45.5) | 112 (12.5) | 0.51 |
| total cholesterol (mg/dL), mean (SD) | 156.7 (38.9) | 164.8 (37.7) | 0.36 |
| HDL(mg/dL), mean (SD) | 53.3 (23.3) | 49.6 (12.7) | 0.28 |
| LDL (mg/dL), mean (SD) | 84.6 (33.7) | 101.5 (77) | 0.32 |
| WBC pre-recanalization<br>(X10<sup>3</sup> /μL) ,mean (SD) | 8.8 (3.6) | 8.7 (3.9) | 0.97 |
| neutrophils pre-recanalization<br>(X10<sup>3</sup> /μL) , mean (SD) | 6.2 (3.3) | 5.6 (3.7) | 0.5 |
| lymphocytes pre-recanalization<br>(X10<sup>3</sup> /μL) , mean (SD) | 1.8 (1.1) | 2.2 (1.1) | 0.18 |
| monocytes pre-recanalization<br>( X10<sup>3</sup> /μL ), mean (SD) | 5.6 (2.2) | 7.3 (6.8) | 0.23 |
| platelets pre-recanalization<br>(X10<sup>3</sup> /μL) , mean (SD) | 221.2 (63.1) | 226 (77.8) | 0.78 |
| SIRI pre-recanalization (X10<sup>3</sup>), mean (SD) | 2.6 (3) | 2.5 (2.8) | 0.79 |
| SII pre-recanalization ( X10<sup>6</sup>), mean (SD) | 1.1 (1.2) | 0.8 (0.8) | 0.13 |
| NLR pre-recanalization, mean (SD) | 5 (5.6) | 3.5 (3.5) | 0.08 |
| NPR pre-recanalization, mean (SD) | 0.03 (0.02) | 0.03 (0.04) | 0.86 |
| **ENDOTHELIAL MARKERS** |  |  |  |
| HArg (μmol/L), mean, (SD) | 1.8 (0.8) | **2.4 (1.2)** | **0.04** |
| SDMA (μmol/L), mean, (SD) | 0.2 (0.2) | 0.2 (0.1) | 0.76 |
| ADMA (μmol/L), mean, (SD) | 0.5 (0.1) | 0.5 (0.1) | 0.45 |
| FMD (μmol/L), median(IQR) | 2(1.2-3) | **3.3(2-6.8)** | **0.05** |
| **STROKE DATA AND ENDOVASCULAR TREATMENT VARIABLES** |  |  |  |
| NIHSS score, median (IQR) | **19 (9.5-22.5)** | 12 (8-18) | **<0.03** |
| cardioembolic vs others, n (%) | 9 (39.1) | 70 (51.1) | 0.37 |
| aspiration devices vs others, n(%) | 13 (59.1) | 84 (69) | 0.37 |
| Time from onset to groin, mean (SD) | 417.3 (284.9) | 316.8 (292.1) | 0.20 |
| door-to-reperfusion time (min), mean (SD) | 194.1 (53.7) | 177.6 (73.9) | 0.39 |
| groin-to-reperfusion time (min), mean (SD) | 63.9 (50.5) | 46.9 (33.3) | 0.07 |
| IVT, n (%) | 4 (17.4) | 43 (31.4) | 0.17 |
| average hospital stay (days), mean (SD) | **11.3 (9.4)** | 8.7 (3.6) | **0.02** |
| **OUTCOMES** |  |  |  |
| GCO at discharge, n (%) | 0 (0) | **62 (49.6)** | **<0.01** |
| GCO at 90 days, n(%) | 1 (4.3) | **75 (57.3)** | **<0.01** |
| futile recanalization, n (%) | **19 (82.6)** | 49 (35.8) | **<0.01** |
| death, n(%) | **12 (52.2)** | 18 (13.1) | **<0.01** |

Abbreviations: sHT: Symptomatic Hemorrhagic transformation group; No-HT: Non-hemorrhagic transformation; SD: standard deviation; AF: atrial fibrillation; LDL: low-density lipoprotein; HDL: high-density lipoprotein; NIHSS: National Institute of Health Stroke Scale; WBC: white blood cells; NLR: neutrophil-to-lymphocyte ratio; NPR: neutrophil-to-platelet ratio; SIRI: systemic inflammation response index; SII: Systemic
Immune-Inflammation Index; HArg: homoarginine; ADMA: asymmetric dimethylarginine; SDMA: symmetric dimethylarginine; FMD: Flow mediated dilation; IVT: intravenous thrombolysis; GCO: good clinical outcome.

**Table 4.** Univariate and multivariate binary regression analysis of factors influencing sHT after stroke treated with EVT.

|  | Univariate OR<br>(95%CI) | p | Adjusted OR<br>(95% CI) | p | Adjusted OR**<br>(95% CI) | p** |
| --- | --- | --- | --- | --- | --- | --- |
| Antiplatelet therapy | 2.62 (1.03-6.68) | 0.04 | 3.05 (0.95-9.72) | 0.06 | 3.04 (0.95-9.74) | 0.06 |
| NIHSS score (per point) | 1.08 (1.01-1.15) | 0.04 | 1.03 (0.95-1.12) | 0.41 | 1.03(0.95-1.12) | 0.42 |
| HArg* | 0.08 (0.01-0.93) | 0.04 | 0.03 (0.02-0.72) | **0.03** | 0.04 (0.02-0.91) | **0.04** |
| FMD* | 0.31 (0.10-0.98) | <0.05 | 0.40 (0.11-1.48) | 0.17 | 0.32 (0.08-1.32) | 0.12 |
| IVT | 0.46 (0.15-1.43) | 0.18 | 0.83 (0.24-2.97) | 0.78 | 0.84 (0.23-3.02) | 0.79 |

Abbreviations: OR: Odds Ratio; NIHSS: National Institute of Health Stroke Scale; HArg:homoarginine; FMD: Flow mediated dilation; IVT: intravenous thrombolysis.
\*: log-transformed
\*\* : Multivariate model including sex, age and variables with a p value < 0.05 in the univariate analysis

## DISCUSSION

Our findings demonstrate that prerecanalization inflammatory markers, including ALC, NLR and SII, independently predicted radiological HT after EVT indicating that an altered inflammatory status during the acute phase of stroke increases the likelihood of this complication. Moreover, endothelial function markers, particularly HArg, correlated with inflammatory parameters such as ANC and WBC, were associated with PH and sHT suggesting that the endothelium derivatives may contribute to more severe phenotypes of HT.

Inflammation plays a central role in the pathophysiological response to ischemic stroke, exerting both deleterious and reparative effects throughout the disease course. Following cerebral ischemia, innate immune cells (microglia, neutrophils, monocytes) and adaptive immune cells (T cells, B cells, and NK cells) participate in a coordinated response that promotes debris clearance and tissue remodeling. However, these same mediators drive acute peri-ischemic injury through proinflammatory signaling and oxidative stress, ultimately promoting blood–brain barrier (BBB) disruption and HT. [22] Neutrophils are a major source of matrix metalloproteinase-9 (MMP-9) in acute ischemic stroke (AIS) and contribute to early BBB damage. [23] Conversely, lymphocyte count is a well-recognized marker of physiologic stress; cortisol-driven lymphopenia has been associated with stress-induced immunosuppression and with exacerbation of ischemic damage through cytokine-mediated pathways. [24,25]

The neutrophil-to-lymphocyte ratio (NLR), which integrates both inflammatory activation and stress-related lymphocyte suppression, has been validated as a robust indicator of inflammatory status in AIS, predicting clinical outcomes [6] and the risk of HT. [17] Consistent with previous evidence, we observed that higher NLR values (1.11 [1.02–1.22], p<0.05) independently increased the risk of HT. The systemic immune-inflammation index (SII), which incorporates platelet count and NLR to capture the interaction between thrombosis and inflammation, has similarly been linked to symptomatic HT after EVT. [26,27] Our results further support this association, showing that elevated prerecanalization SII values confer a greater risk of HT.

Another inflammatory index, SII, combines platelet count and NLR to reflect the interplay between inflammation and thrombosis, two key mechanisms in stroke pathophysiology. SII has also previously been associated with the risk of symptomatic hemorrhagic transformation in stroke patients treated with EVT. [26] [27] In this line, our findings indicate that patients with higher pre-recanalization SII levels were at greater risk of developing HT.

Another pathway involved in peri-ischemic damage and BBB disruption is endothelial dysfunction. Arginine derivatives modulate endothelial homeostasis in cerebrovascular disease: ADMA and SDMA inhibit NO synthase, reducing NO bioavailability and impairing endothelial integrity,[13] whereas HArg serves as a substrate for NO production, promoting endothelial health and vascular homeostasis. [13] Low HArg levels have been linked to endothelial dysfunction and adverse cardiovascular outcomes, including stroke [13]. In the setting of acute ischemic stroke, reduced HArg availability may impair endothelial integrity and regenerative capacity, partly through the diminished function of endothelial progenitor cells (EPC) which play a crucial role in vascular development and reparative responses following vascular injury. Impaired EPC-mediated endothelial recovery exacerbates microvascular instability, facilitating extravasation of blood components into the ischemic parenchyma. Consequently, endothelial dysfunction and insufficient EPC-mediated repair can facilitate hemorrhagic transformation after reperfusion. [28,29]

In our cohort, HArg levels were independently correlated with the radiological PH subtype of HT and with symptomatic HT after reperfusion suggesting that impaired endothelial function, as reflected by HArg, may increase microvascular vulnerability and predispose to more hemorrhagic complications after ischemic stroke.

The observation that HArg but not SDMA or ADMA, associated outcomes after stroke may reflect biological differences in their vascular effects, clearance kinetics, and vascular impacts. While ADMA and SDMA act as systemic markers of endothelial dysfunction, HArg may more directly reflect endothelial NO-producing capacity and BBB preservation, explaining its stronger association with HT. [13] [30]

Moreover, Harg exhibited significant correlations with inflammatory markers, specifically ANC and WBC, whereas SDMA correlated with preALC. The presence of pro-inflammatory markers in the systemic circulation interferes with vascular function and may exert a deleterious effect on NO bioavailability, representing one of the established links between inflammation and the NO pathway in endothelial cells. [11] For instance, elevated CRP levels have been shown to significantly reduce endothelial Nitric Oxide Synthase (eNOS) expression in cultured endothelial cells. [31].

ADMA and SDMA act as mediators of oxidative stress through the inhibition and uncoupling of the NOS enzyme, thereby reducing NO synthesis. On the other hand, inflammation activates arginine methyltransferase proteins (PRMTs) and inhibits DDAHs (dimethylarginine dimethylaminohydrolases), leading to an increase in dimethylarginine, SDMA and ADMA levels[32,33]

In this context, a correlation between SDMA and inflammatory biomarkers such as IL-6 and monocyte chemoattractant protein-1 (MCP-1) has previously been reported [34]. The relationship between HArg and inflammatory biomarkers has not been previously described in the context of stroke, although this molecule may also contribute to oxidative stress.

These findings support the hypothesis that the interaction between inflammation and endothelial integrity not only affects the peripheral vascular bed, but may also exacerbate cerebral ischemic injury, impair collateral perfusion, and limit the efficacy of reperfusion therapies.

Multiple therapeutic approaches under investigation for stroke target inflammatory and NO-endothelium pathways, with varying results. Our study contributes to a more nuanced understanding of the role of the inflammation and NO-endothelium pathways in the context of recanalization therapies and may support the development of better-targeted combination treatment strategies.

This study has several limitations. First, it was a single-center study with a relatively small sample size, which may restrict generalizability of the findings. Second, although we adjusted for relevant confounders, residual confounding cannot be excluded. In this regard and as mentioned in the methods section arginine is a component of the alteplase commercial product, and patients with IVT showed higher mean levels of HArg (2.15 (sd=1.15) vs 2.73 (sd=1.48), p=0.013), however multivariate analysis including IVT did not alter the results of the observed associations.

Third, endothelial biomarkers were measured from arterial samples obtained immediately after groin puncture, whereas inflammatory markers were derived from venous blood drawn earlier, with a mean interval of 40 minutes, which may introduce variability.

Fourth, FMD results should be interpreted cautiously as ultrasound studies were not performed on all patients and all measurements were taken within 72 hours after the stroke event where a loss of cerebrovascular hemodynamic autoregulation is expected, and performed by two different investigators without assessment of interobserver variability.

## CONCLUSION

Taken together, these data support the hypothesis that the interplay between systemic inflammation and endothelial dysfunction significantly influences the risk and severity of HT following EVT. While radiological HT may occur within a broader inflammatory environment, the development of PH and sHT likely requires a combined burden of inflammatory and endothelial impairment.

## DECLARATIONS

### Ethics approval and consent to participate

The study was conducted in accordance with the Declaration of Helsinki and was approved by the Ethics Committee of Gipuzkoa (protocol code PRI-CII-2018-01, approval date February 20, 2018). Written informed consent was obtained from all patients or, when applicable, from their relatives, in accordance with the Spanish Personal Data Protection Law.

### Consent for publication

Not applicable, as this manuscript does not contain any individual patient data, images, or other identifiable information.

### Availability of data and materials

The datasets generated and/or analyzed during the current study are available from the corresponding author on reasonable request.

### Authors’ contributions

Patricia de la Riva designed the study. Kattalin Alvarez, Patricia de la Riva and Jon Rodriguez-Antigüedad collected the clinical data and performed the statistical analysis. Juan Marta-Enguita provided support and advice in the statistical methodology. All authors contributed to drafting and revising the manuscript and approved the final version.

## Data Availability

The datasets generated and/or analyzed during the current study are available from the
corresponding author on reasonable request

## Acknowledgements

The authors would first like to express their gratitude to the patients who participated in this study and to their families for their support. We also extend our sincere thanks to the nursing staff of the angiography unit for their invaluable assistance in collecting and processing blood samples.

