## Supplementary material for "Systemic inflammation and endothelial dysfunction influence the risk and severity of hemorrhagic transformation after endovascular treatment for acute ischemic stroke": Suplementary material

**Supplementary methods**

**Ultrasound assessment of endothelial function, FMD measurement**:
Two investigators (JR during the first year of recruitment and PR during the rest of the study time period) performed ultrasound assessment of systemic endothelial function within 72h after the stroke event by measuring brachial artery FMD in response to reactive hyperemia. The assessment was conducted in keeping with the guidelines of the International Brachial Artery Reactivity Task Force and the Working Group of the European Society of Hypertension [19]. We used a high-resolution B-mode ultrasound (Philips CX50 by Philips ultrasound Andover, MA, USA) with a 7.5-MHz, linear-array transducer. In short, a longitudinal image was used to measure brachial artery diameter (first baseline value, d1). A blood pressure cuff was inflated on the upper arm to 300 mm Hg for 4 min and then deflated. 1 min later, a second longitudinal scan was obtained to calculate the brachial artery diameter (post-occlusion value, d2). FMD was expressed as the percentage of diameter variation (absolute diameter changes were also recorded, FMD = d2 – _d1/d1 x 100). Measurements were repeated twice and median percentage values were obtained for the analysis.

**Supplementary results**

**Table 1.** Post recanalization inflammatory markers in patients with HT compared to No-TH patients.

| | **HT** | **No-HT** | **p** |
|---|---|---|---|
| n (%) | 23 (7.7) | 137 (59.8) | |
| **INFLAMMATORY MARKERS** | | | |
| WBC post-recanalization ($X10^3$ /µL) ,mean (SD) | 10.1 (3.2) | 9.7 (4.1) | 0.43 |
| neutrophils post-recanalization ($X10^3$ /µL) , mean (SD) | 7.8 (3.0) | 7.2 (4.1) | 0.25 |
| lymphocytes post-recanalization ($X10^3$ /µL) , mean (SD) | 1.3 (0.5) | 1.4(0.6) | 0.06 |
| monocytes post-recanalization ( $X10^3$ /µL ), mean (SD) | 0.7 (0.3) | 0.7(0.3) | 0.87 |
| platelets post-recanalization ($X10^3$ /µL) , mean (SD) | 195.8 (69.7) | 201.7 (61.7) | 0.60 |
| CRP post-recanalization (mg/L) mean (SD) | 40.5(56.5) | 38.5(60.2) | 0.84 |
| SIRI post-recanalization ($X10^3$ ), mean (SD) | 5.5 (4.2) | 5.0 (7.1) | 0.59 |
| SII post-recanalization ( $X10^6$), mean (SD) | 1.3 (0.9) | 1.3 (1.6) | 0.82 |
| NLR post-recanalization, mean (SD) | 5 (5.6) | 3.5 (3.5) | 0.08 |
| NPR post-recanalization, mean (SD) | 0.03 (0.02) | 0.03 (0.04) | 0.86 |

Abbreviations: HT: hemorrhagic transformation; No-HT: Non-hemorrhagic transformation; SD: standard deviation; WBC: white blood cells; NLR: neutrophil-to-lymphocyte ratio; NPR: neutrophil-to-platelet ratio; CRP: C-reactive protein; SIRI: systemic inflammation response index; SII: Systemic Immune-Inflammation Index.
